# The real-world effectiveness of an intranasal spray A8G6 antibody cocktail in the post-exposure prophylaxis of COVID-19

**DOI:** 10.1101/2023.03.14.23287255

**Authors:** Xiaosong Li, Pai Peng, Haijun Deng, Qian Yang, Shi Chen, Benhua Li, Miao He, Zhu Yang, Ni Tang, Ailong Huang

## Abstract

**Background:** Due to the continuous appearance of novel SARS-CoV-2 variants that are resistant to approved antibodies and leading to the epidemic rebound, several approved neutralizing antibodies have been paused for their usage against COVID-19. Previously, we identified A8G6, an antibody combination of two synergic SARS-CoV-2 neutralizing antibodies 55A8 and 58G6, that showed broad neutralizing activities against Omicron variants. When administrated by the nasal spray delivery system, A8G6 showed promising efficacy in COVID-19 animal models and also showed favorable safety profile in preclinical models as well as in a first-in-human trial. The aim of this study is to evaluate the real-world efficacy of A8G6 neutralizing antibody nasal spray in post-exposure prevention of COVID-19.

**Methods:** From November 27, 2022 to January 31, 2023, an open-label, non-randomized, two-arm, blank-controlled, investigator-initiated trial was conducted in Chongqing, China. High-risk healthy participants (18-65 years) within 72 hours after close contact to SARS-CoV-2 infected individuals were recruited and received a three-dose (1.4 mg/dose) A8G6 nasal spray treatment daily or no treatment (blank control) for 7 consecutive days. The primary end points were 1) the occurrence of positive SARS-CoV-2 RT-PCR cases in A8G6 treated group vs blank control group at the end of day 7; 2) time to SARS-CoV-2 positive conversion at the end of day 7. The secondary end points were 1) viral load of SARS-CoV-2 when participants became SARS-CoV-2 positive; 2) the time from SARS-CoV-2 infection to negative COVID-19 conversion. Safety end point of the nasal spray AG86 was analyzed by recording adverse events during the whole course of this trial. This study was registered with Chictr.org (ChiCTR2200066416).

**Findings:** Of 513 enrolled participants, 173 in the A8G6 treatment group and 340 in the blank-control group were included in the analysis. SARS-CoV-2 infection occurred in 151/340 (44.4%) subjects in the blank control group and 12/173 (6.9%) subjects with the A8G6 treatment group. The result indicates that the intranasal spray A8G6 reduces the risk of SARS-CoV-2 infection (HR=0.12, 95% CI, 0.07-0.22; *p*<0.001). The prevention efficacy of the A8G6 treatment within 72-hours exposure was calculated to be 84.4% (95% CI: 74.4%-90.4%). Moreover, compared to the blank-control group, the time from the SARS-CoV-2 negative to the positive COVID-19 conversion was significantly longer in the AG86 treatment group (mean time: 3.4 days in the A8G6 treatment group vs 2.6 days in the control group, *p*=0.019). In the secondary end-point analysis, the A8G6 nasal treatment had no effects on the viral load at baseline SARS-CoV-2 RT-PCR positivity and the time of the negative COVID-19 conversion (viral clearance). Finally, 5 participants (3.1%) in the treatment group reported general adverse effects. We did not observe any severe adverse effects related to the A8G6 treatment in this study.

**Interpretation:** In this study, the intranasal spray AG86 antibody cocktail showed potent efficacy for prevention of SARS-CoV-2 infection in close contacts of COVID-19 patients.

**Funding:** Chongqing Biomedical R&D Major Special Project, Project (No. CSTB2022TIAD-STX0013), Chongqing Science and Health Joint Medical High-end Talent Project (No. 2022GDRC012), Science and Technology Research Program of Chongqing Municipal Education Commission (No. KJZD-K202100402), CQMU Program for Youth Innovation in Future Medicine (No. W0073).

**Research in context:** *Evidence before the study:* Two potent neutralizing antibodies 55A8 and 58G6 against SARS-CoV-2 were identified from the plasma of COVID-19 convalescent patients. In our previous studies, the synergetic neutralization of the antibody combination of 55A8 and 58G6 (A8G6) had been shown in structural mechanism, as well as in vitro and in vivo. Pre-clinical evaluation of A8G6 nasal spray showed promising efficacy against Omicron BA.4/5 infection in golden syrian hamsters challenged with live virus. In a first-in-human trial, A8G6 also showed favorable safety profile and nasal concentration over IC90 of neutralization activity against Omicron BA.4/5. The preliminary data showed that the intranasal spray A8G6 had the excellent efficacy, safety and druggability to protect against COVID-19.

*Added value of this study:* This is the first human trial showing that a nasal spray of neutralizing antibody cocktail is efficacious in preventing SARS-CoV-2 infection but is not efficacious in the post-infection treatment of COVID-19. In the Omicron wave of the COVID-19 pandemic in China in November, 2022, COVID-19 close contacts receiving the A8G6 treatment in the designated quarantine hotels showed a significantly lower incidence of SARS-CoV-2 infection. Additionally, the A8G6 treatment delayed time from exposure to the diagnosis of the COVID-19 positivity (median time: 3.4 days in the treatment group vs 2.6 days in the control group). Furthermore, we analyzed the effects of the A8G6 treatment on the clinical status of close contacts who became infected with SARS-CoV-2. Results suggests that there were no significant differences in viral load of SARS-CoV-2 at the beginning of positive infection and the time of the viral clearance between A8G6 treatment and blank control groups. Overall, the trial result is consistent with the mechanism of action of nasal spray antibody cocktail for the prevention of SARS-CoV-2 infection. Finally, low safety risk of the nasal spray A8G6 was also shown in the trial.

*Implications of all the available evidence:* We observed the use of A8G6 to reduce the risk of SARS-CoV-2 infection. This study provided supporting evidences for the real-world effectiveness and safety of the nasal spray A8G6 among high-risk close contacts in the post-exposure prevention of COVID-19 during the Omicron BA.5.2 wave in China. This is the first proof of concept of using nasal spray neutralizing antibody for the prevention of viral infection. It implicates that the promising efficacy of the nasal spray A8G6 makes it possible for the fast-acting prevention in future COVID-19 waves.

## 1. Introduction

At present, SARS-CoV-2 is still defined as a Public Health Emergency of International Concern. Due to the continuous evolution of SARS-CoV-2, its variants led to a high risk of COVID-19 global transmission. Although vaccination played important roles in the preventing and controlling of COVID-19, ^1, 2^ the neutralizing antibodies (NAbs) elicited by vaccines were heterogeneous among different individuals and were waning within several months. ^3–5^ NAbs blocking the entry of SARS-CoV-2 into host cells have been developed for the COVID-19 prevention or therapy. Several SARS-CoV-2 targeting monoclonal antibodies (mAbs) have previously been authorized for use through an emergency use authorization (EUA).^6–10^ However, due to the failure or significant decrease of neutralization against some emerging SARS-CoV-2 variants, the usage of these antibody drugs was limited. There is an urgent need to develop broad-spectrum and effective NAbs against the circulating and other novel SARS-CoV-2 variants. Furthermore, those approved neutralizing antibodies, when administrated systematically, provided limited efficacy in the prevention of viral infection. We hypothesized that this was due to the low concentration of those neutralizing antibodies at nasal compartment when administered systematically. As a potentially more effective prophylactic approach, we proposed to use neutralizing antibodies as nasal spray to prevent viral infection at the viral entry point to human body.

A8G6 was a combination of 58G6 and 55A8 monoclonal NAbs which were identified from COVID-19 convalescent patients at early 2020.^11^ Previous study showed that 58G6 recognized both the steric site S^470–495^ and another region S^450-458^ on the receptor binding domain (RBD) of SARS-CoV-2 spike protein (S protein). When administrated as a nasal spray, 58G6 demonstrated prophylactic efficacy against authentic SARS-CoV-2 ancestral strain and the Beta variant (B.1.351) in the transgenic mice expressing human ACE2 (hACE2) and against Delta and Omicron variants in hamster model.^12, 13^ 55A8 exhibited potent binding affinities to the S proteins of ancestral SARS-CoV-2 strain, Delta, Omicron BA.1, BA.2, and BA.4/5 at sub-picomolar level. When the two NAbs simultaneously interacted with S protein, 58G6 and 55A8 recognized different and complementary epitopes in RBD of SARS-CoV-2 and further occluded the accessibility of the S protein to ACE2. Therefore, A8G6 antibody cocktail which consisted of two potent neutralizers 58G6 and 55A8 displayed a synergetic potency and the broad neutralization against the Omicron variants.^14^ Moreover, intranasal delivery of the cocktail A8G6 demonstrated potent protection against Omicron in hamster model. A first-in-human trial of the intranasal spray A8G6 antibody cocktail in healthy volunteers provided evidences for safety and the potential clinical efficacy in preventing Omicron BA.4/5 infections (unpublished, manuscript in preparation). The real-world effectiveness of the A8G6 nasal spray needs to be further evaluated.

Here we conducted an open-label, non-randomized, two-arm, blank-controlled trial among close contacts of COVID-19 patients in several designated quarantine hotels to assessed the effectiveness and safety of A8G6 intranasal spray for the post-exposure prophylaxis of COVID-19 during the Omicron BA.5.2 wave occurred in November, 2022 in Chongqing, China.

## 2. Methods

### Study design

In this study, an open-label, non-randomized, two-arm, blank-controlled, investigator-initiated trial was designed to assess the efficacy and safety of the intranasal spray cocktail A8G6 in preventing SARS-CoV-2 infection among close contacts with COVID-19 patients. The clinical trial was conducted at 6 designated quarantine hotels in Yuzhong District, Chongqing, China from November 27, 2022 and was completed on December 12, 2023.

Recruited participants in the treatment group self-administrated a three doses of 0.7 mg (140μl) A8G6 nasal spray per day for 7 treatment days. The drug was supplied by Chongqing Mingdao Haoyue Biotechnology Co., LTD (Chongqing, China), stored at 2-8 °C. In the blank control group, participants did not receive any treatment. After enrollment, SARS-CoV-2 infection was confirmed by a reverse transcription polymerase chain reaction (RT-PCR) test of oropharyngeal swab. During this trial, with the adaption of the anti-COVID-19 policy, not only RT-PCR, but also rapid antigen tests were used to confirm the SARS-CoV-2 infection status.

The trial was carried out in accordance with all applicable national and local regulatory requirements. Data and Safety Monitoring Board of The Second Affiliated Hospital of Chongqing Medical University oversaw trial conduct and documentation. The protocol has been approved by the Chinese clinical test registration center (the world health organization international clinical trials registered organization registered platform (ICTRP), the registration number: ChiCTR2200066416) and the Ethics Committees of The Second Affiliated Hospital of Chongqing Medical University (the approval number: 2022127-1).

### Participants

During November COVID-19 wave in Chongqing, China, when patients had been diagnosed as COVID-19 with the positive RT-PCR test for SARS-CoV-2 (index cases), their close contacts were immediately transferred to the designated quarantine sites. At 6 quarantine sites in Chongqing, healthy adults aged between 18 to 65 years who had a close contact with index cases within 72 hours were enrolled into this study. The maximum time interval between exposure to treatment was ≤ 72 hours. All vaccination status is eligible for inclusion. Exclusion criteria included positive RT-PCR at baseline, nasal discomfort, the use of other COVID-19 antibody drugs and drug-drug interference with participants’ regular medication (additional details about eligibility criteria were described in the appendix).

All study participants were capable of self-administrating the intranasal spray, recording and recalling clinical signs. All participants were provided and voluntarily signed written informed consent before the study.

### Procedures

At six quarantine sites in the Yuzhong District, Chongqing, site investigation was carried out to screen eligible participants. Eligible participants were given the choice to join the A8G6 treatment group or blank control group. For eligible participants that showed “no preference” in either group, they were randomly assigned to A8G6 treatment group or blank control group. Oropharyngeal swabs were taken for quantitative and qualitative RT-PCR assessments at baseline prior to treatment and though the treatment period and a follow-up period. Subjects with positive RT-PCR results before treatment were excluded. The SARS-CoV-2 viral load was present by viral genome copies per mL log10 values with the conversion of the open reading frame of 1ab (ORF1ab) and nucleocapsid (N-gene) cycle threshold (Ct) values (RT-PCR was conducted by Yuzhong District Center for Disease Control and Prevention, in Chongqing, China. Conversion of Ct values to viral genome copies was calculated according to the manufacturer’s instructions of 2019-nCoV viral RNA kit produced by BioPerfectus Technologies, catalog number: JC10223-1N).

Subjects’ demographic data, health and COVID-19 vaccination status were recorded at the baseline visit (Day 0). The use of nasal spray, rapid antigen tests or RT-PCR test for COVID-19 were recorded every day during the study participation. When participants in both groups were diagnosed with SARS-CoV-2 infection, the related symptoms and symptomatic treatment for COVID-19 were reported until the trial completed. In the treatment group, all participants were requested to self-report and record the adverse events. Due to the relaxation of COVID-19 control and policy starting from December 4, 2022, some participants returned to home for further isolation. The follow-up visits were adjusted to retrospective telephonic visit according to a questionnaire form from that day.

### Outcomes

The primary endpoint analysis included all participants in both the treatment and control groups. The primary endpoint was to assess the efficacy of the intranasal spray A8G6 for post-exposure prophylaxis of COVID-19. In this study, we compared the COVID-19 incidence of the close contacts between the A8G6 treatment individuals and the blank-controlled individuals. We also compared the time from enrollment to SARS-CoV-2 infection between the two groups. The secondary efficacy analysis included the quantitative data of SARS-CoV-2 RNA (log10 copies per mL) at baseline of the positive COVID-19, the time to conversion of SARS-CoV-2 RNA from positive to negative (viral clearance) and the time to symptom remission of COVID-19 patients.

Safety endpoints was adverse event types and the incidence rate of adverse events among all participants of the A8G6 treatment group during the study. An adverse effect was defined as any abnormal signs or symptoms and harmful results caused by the study drug.

### Statistical analysis

The sample size in this clinical trial was determined on the basis of statistical power calculations. We proposed greater than 90% power to detect a 20% relative difference between the A8G6 treated and control group at a two-sided alpha level of 0.05 (ie., a 20% prevention efficacy of A8G6). The formula is as follows:

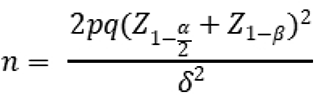

which p is the proportion of participants develop COVID-19 in A8G6 treated group, q is in the control group, δ is the difference between two group, α is two-sided alpha level, and 1-β is statistical power. In this clinical trial, we assume that q is 0.1, 20% relative reduction of A8G6 treated group is 0.08. Assuming a dropout rate of 20%, at total of 5160 participants will be recruited.

The primary efficacy endpoints including COVID-19 incidence and time to confirmed SARS-CoV-2 infection. The COVID-19 incidence was analyzed using the Kaplan-Meier method and log-rank test, and the time to confirmed SARS-CoV-2 infection was analyzed using Wilcoxon rank-sum test. The secondary efficacy endpoints including viral load when confirmed SARS-CoV-2 infection and the time to negative conversion of SARS-CoV-2 determined by RT-PCR. The viral load when confirmed SARS-CoV-2 infection was analyzed using Wilcoxon rank-sum test, negative conversion of SARS-CoV-2 and remission time were conducted using Kaplan-Meier method and log-rank-test. Safety was assessed in participants in the full analysis set who received A8G6 nasal spray treatment during the 8-day quarantine period.

Database from the Service Platform for COVID-19 Prevention and Control created by Yuzhong District Center for Disease Control and Prevention were authorized for us to use and analyze. Data including demographic and clinical characteristics of the cohorts, endpoints in this clinical trial were collected from an applet of WeChat (a social media platform in China), called “Yuzhong Information Exchange”. All data were summarized with descriptive statistics (number of subjects (%), median (IQR), mean±sd). The credible interval for nasal spray was calculated with the use of a beta-binomial model with prior beta (1, 1) adjusted for the treatment duration time. Continuous variables were compared with the Mann–Whitney U-test, and Categorical variables were conducted using χ^2^ test or Fisher’s exact test. A P value of <0.05 was considered statistically significant. Statistical analyses were performed using R software, version 3.6.0.

### Role of the funding source

The funder of the study had no role in study design, data collection, data analysis, data interpretation, or writing of the report. All authors had full access to all the data in the study and had final responsibility for the decision to submit for publication.

## 3. Results

Since November 27, 2022, a total of 657 individuals were screened in the designated quarantine hotels. There were 101 individuals excluded according to the inclusion and exclusion criteria. The remaining 556 individuals were assigned into either A8G6 treatment group or blank-controlled group based on their preference during singing of consent form. For participants who indicated “no preference” in study group assignment, they were randomly assigned to A8G6 treatment group or the blank control group. Ten participants in the treatment group and 33 participants in the control group were excluded due to consent withdrawal or loss to follow up (Figure 1). The full analysis set (n=513) included all participants who received the A8G6 treatment or blank-control and completed the study. The per-protocol population (n=162) in the treatment group comprised participants who received the A8G6 treatment or no treatment, were treated within 72 hours after exposure.

**Figure 1.**
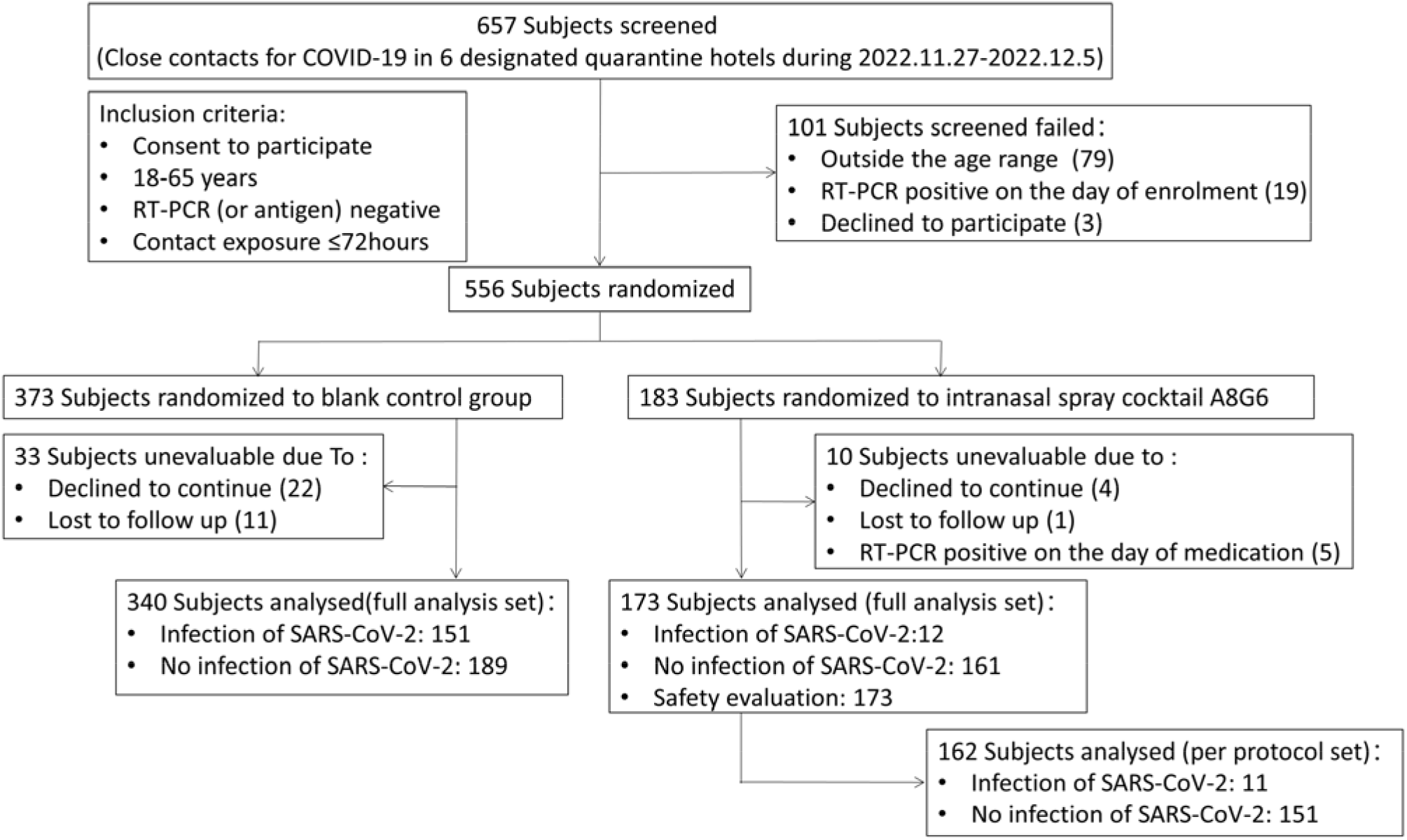
Screening and follow-up of participants.

The final number of subjects completing the trial was 173 subjects in the A8G6 treatment group and 340 subjects in the control group, respectively. In the treatment group, 4 participants started to self-administrated A8G6 at the same day after exposure (Day 0); 73 participants used the nasal spray at the first day after exposure (Day 1); 49 participants at the second days after exposure (Day 3); 35 participants at the third day after exposure (Day 3) and 12 participants at more than 4 days after exposure (Day≥4). Among all participants in the full analysis set, median age was 36.0 (interquartile range, IQR: 26.0-48.0) years; there was a comparable sex ratio between the A8G6 group (55.5% for male and 44.5% for female) and the control group (58.2% for male and 41.8% for female); median BMI was 22.9 (IQR: 20.8-25.4); 18 (10.4%) participants in the treatment group have comorbidities, while 44 (12.9%) participants in the control group have comorbidities. 98.6% participants received different doses of COVID-19 vaccines (Table 1).

**Table 1.**
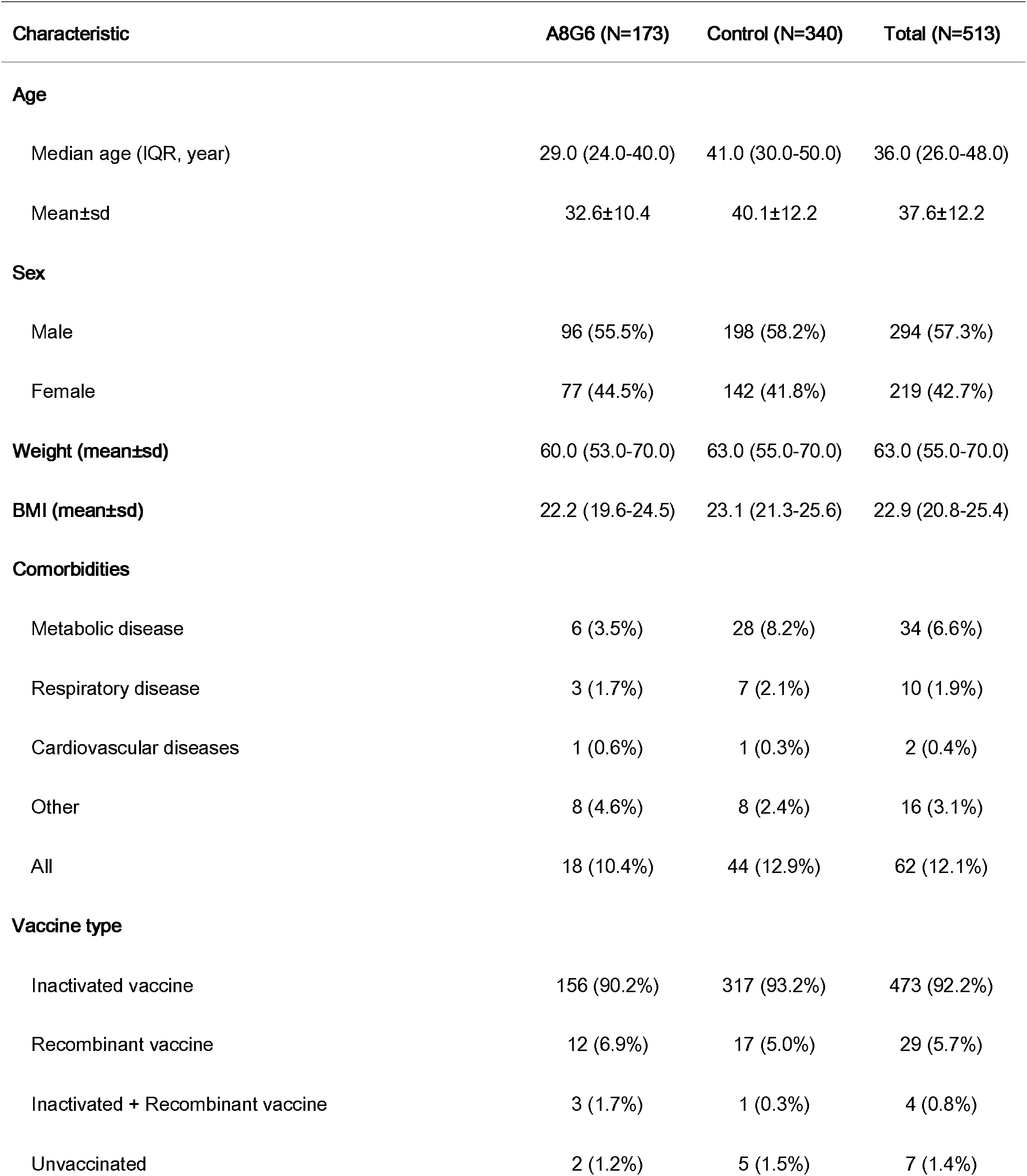
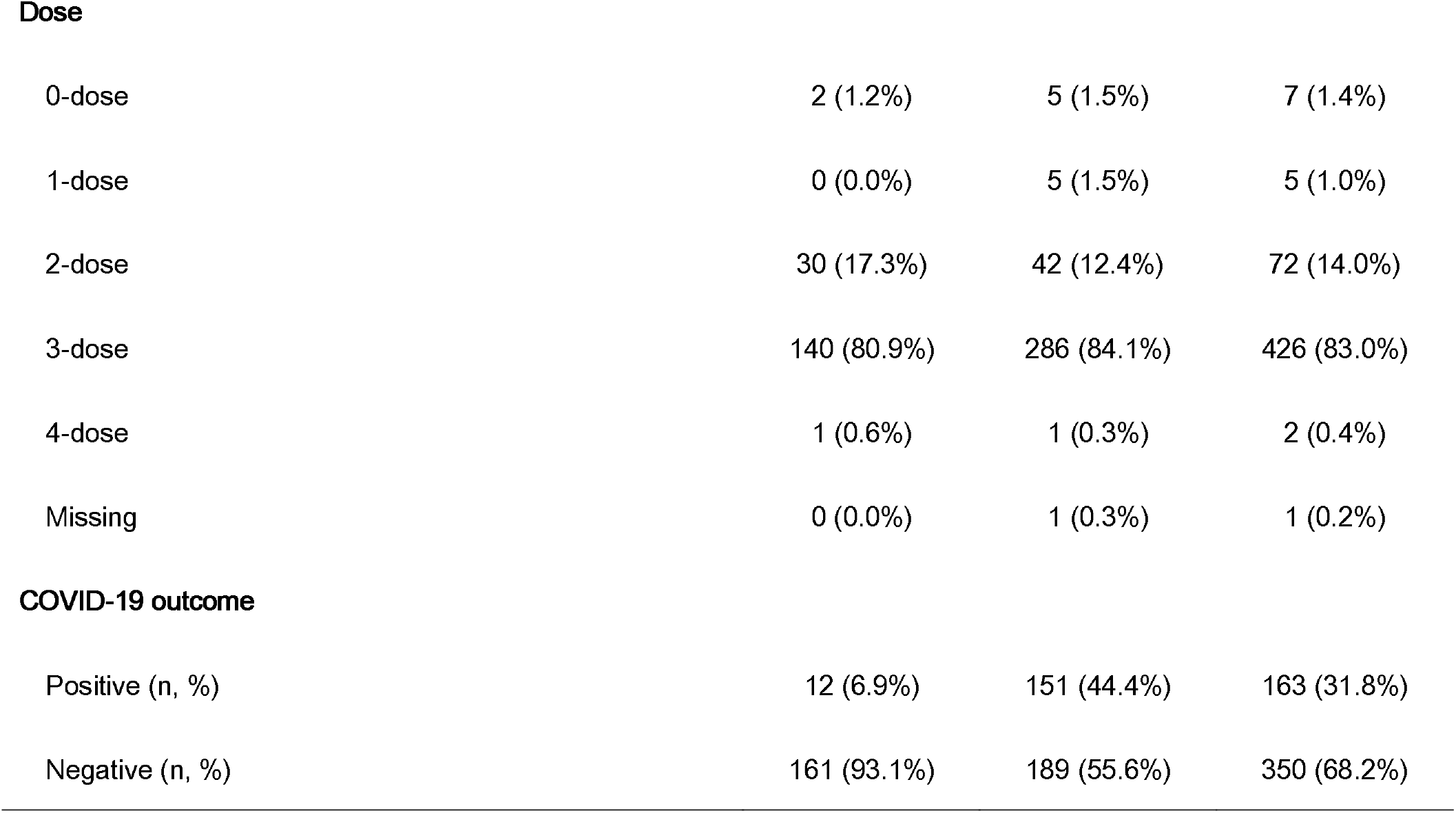
Demographic and Clinical Characteristics of the Cohort.

| Characteristic | A8G6 (N=173) | Control (N=340) | Total (N=513) |
| --- | --- | --- | --- |
| **Age** |  |  |  |
| Median age (IQR, year) | 29.0 (24.0-40.0) | 41.0 (30.0-50.0) | 36.0 (26.0-48.0) |
| Mean±sd | 32.6±10.4 | 40.1±12.2 | 37.6±12.2 |
| **Sex** |  |  |  |
| Male | 96 (55.5%) | 198 (58.2%) | 294 (57.3%) |
| Female | 77 (44.5%) | 142 (41.8%) | 219 (42.7%) |
| **Weight (mean±sd)** | 60.0 (53.0-70.0) | 63.0 (55.0-70.0) | 63.0 (55.0-70.0) |
| **BMI (mean±sd)** | 22.2 (19.6-24.5) | 23.1 (21.3-25.6) | 22.9 (20.8-25.4) |
| **Comorbidities** |  |  |  |
| Metabolic disease | 6 (3.5%) | 28 (8.2%) | 34 (6.6%) |
| Respiratory disease | 3 (1.7%) | 7 (2.1%) | 10 (1.9%) |
| Cardiovascular diseases | 1 (0.6%) | 1 (0.3%) | 2 (0.4%) |
| Other | 8 (4.6%) | 8 (2.4%) | 16 (3.1%) |
| All | 18 (10.4%) | 44 (12.9%) | 62 (12.1%) |
| **Vaccine type** |  |  |  |
| Inactivated vaccine | 156 (90.2%) | 317 (93.2%) | 473 (92.2%) |
| Recombinant vaccine | 12 (6.9%) | 17 (5.0%) | 29 (5.7%) |
| Inactivated + Recombinant vaccine | 3 (1.7%) | 1 (0.3%) | 4 (0.8%) |
| Unvaccinated | 2 (1.2%) | 5 (1.5%) | 7 (1.4%) |
| **Dose** |  |  |  |
| 0-dose | 2 (1.2%) | 5 (1.5%) | 7 (1.4%) |
| 1-dose | 0 (0.0%) | 5 (1.5%) | 5 (1.0%) |
| 2-dose | 30 (17.3%) | 42 (12.4%) | 72 (14.0%) |
| 3-dose | 140 (80.9%) | 286 (84.1%) | 426 (83.0%) |
| 4-dose | 1 (0.6%) | 1 (0.3%) | 2 (0.4%) |
| Missing | 0 (0.0%) | 1 (0.3%) | 1 (0.2%) |
| **COVID-19 outcome** |  |  |  |
| Positive (n, %) | 12 (6.9%) | 151 (44.4%) | 163 (31.8%) |
| Negative (n, %) | 161 (93.1%) | 189 (55.6%) | 350 (68.2%) |

**Table 2.** Demographic and Clinical Characteristics of COVID-19 positive individuals.

| Characteristic | A8G6 (N=12) | Control (N=151) | P value |
| --- | --- | --- | --- |
| **Age** |  |  | 0.103 |
| Median age (IQR, year) | 36.0 (31.2-45.5) | 42.0 (32.0-51.0) |  |
| Mean±sd | 36.3±10.4 | 41.8±12.0 |  |
| **Sex** |  |  | 0.375 |
| Male | 8 (66.7%) | 77 (51.0%) |  |
| Female | 4 (33.3%) | 74 (49.0%) |  |
| **Weight (mean±sd)** | 61.5 (58.8-66.2) | 63.0 (55.0-70.0) | 0.934 |
| **BMI (mean±sd)** | 21.4 (20.9-23.2) | 23.7 (21.4-26.0) | 0.057 |
| **Clinical phenotype** |  |  | 0.694 |
| Symptomatic | 11 (91.7%) | 127 (84.1%) |  |
| Asymptomatic | 1 (8.3%) | 24 (15.9%) |  |
| **Days to COVID-19 confirmed** |  |  | 0.019 |
| median days, IQR | 3.5 (2.8-4.0) | 3.0 (2.0-3.0) |  |
| Mean±sd | 3.4±1.1 | 2.6±1.2 |  |
| **Duration of SARS-CoV-2 positive (day)** |  |  | 0.724 |
| median days, IQR | 6.5 (5.0-7.2) | 7.0 (4.0-7.0) |  |
| Mean±sd | 6.7±1.9 | 6.3±2.5 |  |
| **Viral load (Conversion according to Ct value**<br>**of ORF1ab gene)** |  |  | 1.000 |
| High level, >10<sup>5</sup> copies/ml | 5 (41.7%) | 69 (45.7%) |  |
| Low level, <10<sup>5</sup> copies/ml | 7 (58.3%) | 82 (54.3%) |  |
| **Viral load (Conversion according to Ct value of N gene)** |  |  | 0.117 |
| High level, >10<sup>5</sup> copies/ml | 5 (41.7%) | 100 (66.2%) |  |
| Low level, <10<sup>5</sup> copies/ml | 7 (58.3%) | 51 (33.8%) |  |
| **Symptomatic treatment** |  |  | 0.039 |
| Western medicine | 11 (91.7%) | 89 (58.9%) |  |
| Traditional chinese medicine | 0 (0.0%) | 8 (5.3%) |  |
| Combination of western and traditional chinese medicine | 1 (8.3%) | 6 (4.0%) |  |
| Untreated | 0 (0.0%) | 48 (31.8%) |  |
| **Duration of Covid-19 symptoms (d)** |  |  | 0.401 |
| median days, IQR | 5.0 (1.5-5.0) | 3.0 (2.0-4.5) |  |
| Mean±sd | 4.3±2.9 | 4.2±5.0 |  |
| **Comorbidities** |  |  | 0.435 |
| Metabolic disease | 1 (8.3%) | 17 (11.3%) |  |
| Respiratory disease | 1 (8.3%) | 2 (1.3%) |  |
| Cardiovascular diseases | 0 (0.0%) | 1 (0.7%) |  |
| Other | 1 (8.3%) | 5 (3.3%) |  |
| All | 3 (25.0%) | 25 (16.6%) |  |
| **Vaccine type** |  |  | 0.203 |
| Inactivated vaccine | 10 (83.3%) | 143 (94.7%) |  |
| Recombinant vaccine | 2 (16.7%) | 5 (3.3%) |  |
| Unvaccinated | 0 (0.0%) | 3 (2.0%) |  |
| **Dose** |  |  | 1.000 |
| 0-dose | 0 (0.0%) | 3 (2.0%) |  |
| 1-dose | 0 (0.0%) | 1 (0.7%) |  |
| 2-dose | 2 (16.7%) | 24 (15.9%) |  |
| 3-dose | 10 (83.3%) | 123 (81.5%) |  |
| **Signs and symptoms** |  |  |  |
| Fever | 10 (83.3%) | 83 (55.0%) | 0.071 |
| Fatigue | 4 (33.3%) | 22 (14.6%) | 0.102 |
| Dry cough | 6 (50.0%) | 54 (35.8%) | 0.361 |
| Headache | 3 (25.0%) | 39 (25.8%) | 1.000 |
| Dizziness | 0 (0.0%) | 5 (3.3%) | 1.000 |
| Ageusia | 2 (16.7%) | 8 (5.3%) | 0.161 |
| Pharyngalgia | 1 (8.3%) | 12 (7.9%) | 1.000 |
| Myalgia | 2 (16.7%) | 37 (24.5%) | 0.733 |
| Chill | 1 (8.3%) | 3 (2.0%) | 0.266 |
| Rhinorrhea | 0 (0.0%) | 2 (1.3%) | 1.000 |
| Nasal congestion | 0 (0.0%) | 2 (1.3%) | 1.000 |
| Diarrhea | 0 (0.0%) | 4 (2.6%) | 1.000 |
| Anorexia | 1 (8.3%) | 6 (4.0%) | 0.421 |
| Vomiting | 1 (8.3%) | 1 (0.7%) | 0.142 |
| Arthralgia | 2 (16.7%) | 4 (2.6%) | 0.063 |
| Nausea | 0 (0.0%) | 2 (1.3%) | 1.000 |
| Abdominal pain | 0 (0.0%) | 1 (0.7%) | 1.000 |
| Dry throat | 0 (0.0%) | 2 (1.3%) | 1.000 |
| Insomnia | 0 (0.0%) | 2 (1.3%) | 1.000 |
| Somnolence | 0 (0.0%) | 2 (1.3%) | 1.000 |
| Asthma | 0 (0.0%) | 1 (0.7%) | 1.000 |
| Expectoration | 1 (8.3%) | 0 (0.0%) | 0.074 |
| Eye swelling | 1 (8.3%) | 0 (0.0%) | 0.074 |
| Any | 11 (91.7%) | 127 (84.1%) | 0.694 |

### Efficacy of A8G6 nasal spray in the post-exposure prevention of SARS-CoV-2 infection

After enrollment, oropharyngeal swabs of all subjects in the full analysis set were taken for RT-PCR test for SARS-CoV-2 infection every day. In total, 163/513 (31.8%) participants developed COVID-19 during the 14-day follow-up study. Among them, 12/173 (6.9%) individuals were in the A8G6 treatment group and 151/340 (44.4%) were in the blank control group (Table 1-3, Figure 2A). This difference in COVID-19 incidence rate between groups was statistically significant (Hazard ratio, HR=0.12, 95% CI, 0.07-0.22; log-rank *p*<0.001). The mean (±SD) time of the positive COVID-19 conversion was significantly longer in the A8G6 group compared to the control group (3.4±1.1 days vs 2.6±1.2 days, p=0.019) (Figure 2B). Similar results of data analysis were obtained in the per protocol set (data not shown).

**Figure 2.**
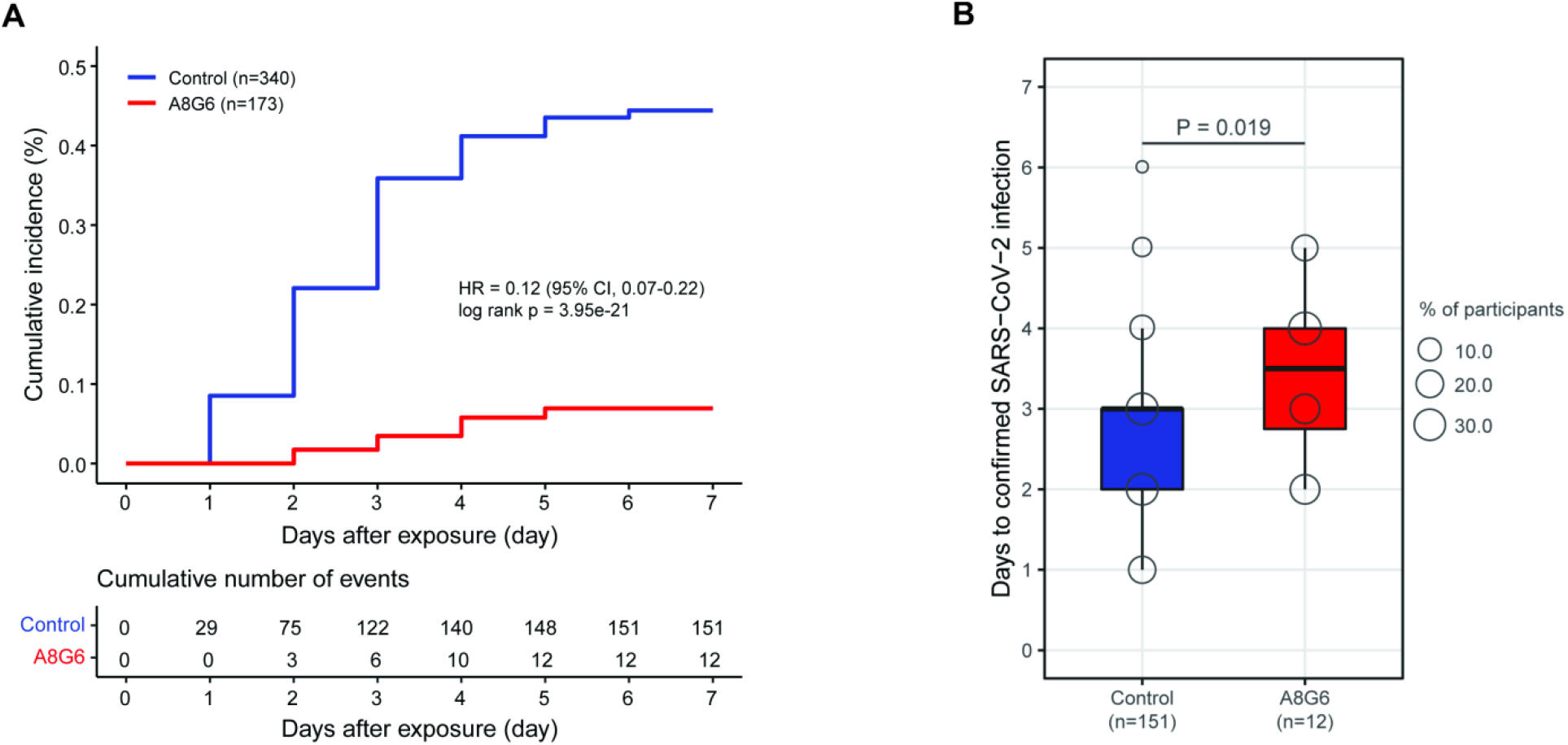
Kaplan Meier plot of occurrence of RT-PCR-confirmed COVID-19.

### The effect of A8G6 on the viral load of SARS-CoV-2 infection at baseline

After enrollment, oropharyngeal swabs of all subjects were taken for RT-PCR test for SARS-CoV-2 every day. When participants were diagnosed as SARS-CoV-2 infection, the Ct values of ORF1ab and N genes were recorded and converted into copies per mL log10 values. Five subjects (41.7%) in the A8G6 treatment group had high viral load (>10^5^ copies/ml) of the ORF1ab gene, compared with 69 subjects (45.7%) in the control group (Table 3); Five subjects (41.7%) in the A8G6 treatment group had high viral load of the N gene (>10^5^ copies/ml), compared with 100 subjects (66.2%) in the control group.

**Table 3.**
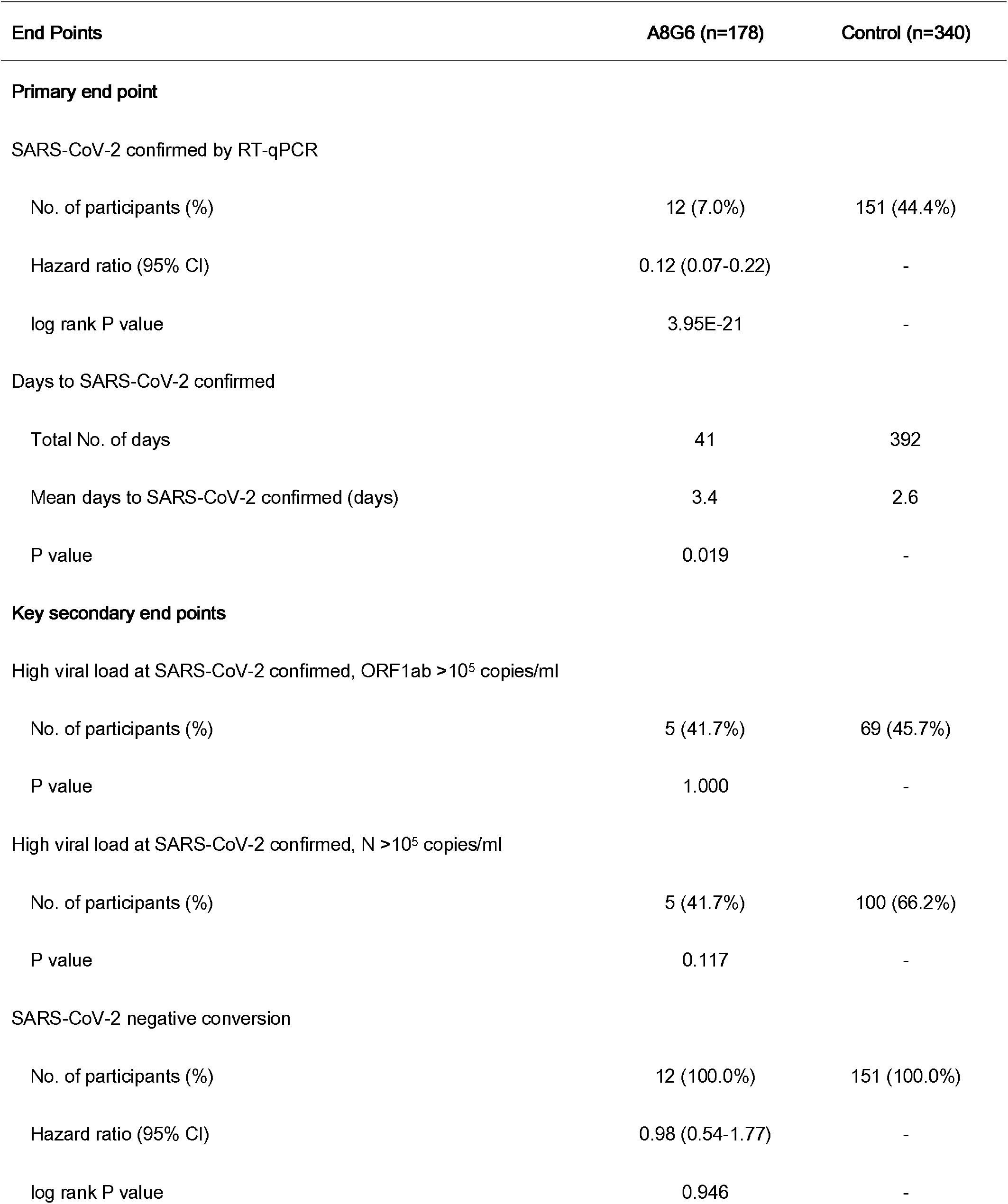
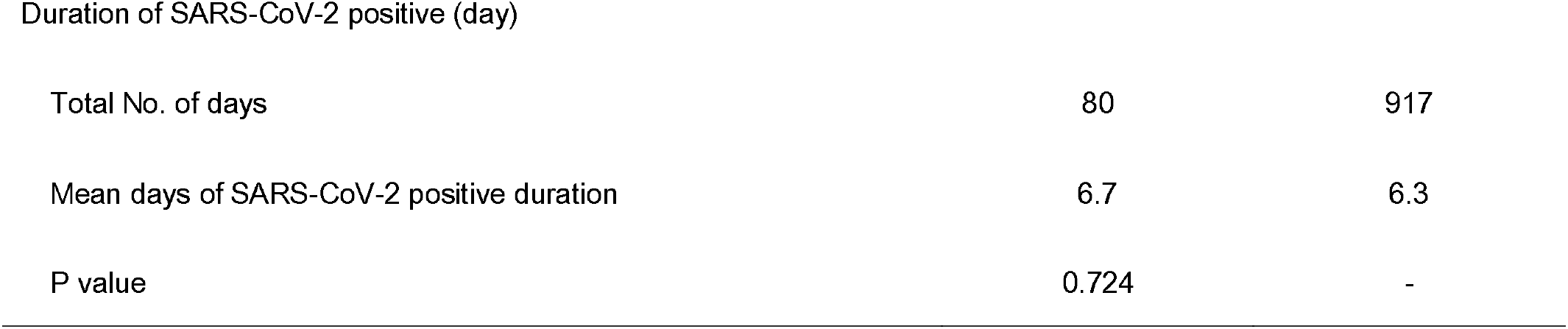
Primary and Key Secondary Efficacy End Points.

| End Points | A8G6 (n=178) | Control (n=340) |
| --- | --- | --- |
| **Primary end point** |  |  |
| SARS-CoV-2 confirmed by RT-qPCR |  |  |
| No. of participants (%) | 12 (7.0%) | 151 (44.4%) |
| Hazard ratio (95% CI) | 0.12 (0.07-0.22) | - |
| log rank P value | 3.95E-21 | - |
| Days to SARS-CoV-2 confirmed |  |  |
| Total No. of days | 41 | 392 |
| Mean days to SARS-CoV-2 confirmed (days) | 3.4 | 2.6 |
| P value | 0.019 | - |
| **Key secondary end points** |  |  |
| High viral load at SARS-CoV-2 confirmed, ORF1ab >10<sup>5</sup> copies/ml |  |  |
| No. of participants (%) | 5 (41.7%) | 69 (45.7%) |
| P value | 1.000 | - |
| High viral load at SARS-CoV-2 confirmed, N >10<sup>5</sup> copies/ml |  |  |
| No. of participants (%) | 5 (41.7%) | 100 (66.2%) |
| P value | 0.117 | - |
| SARS-CoV-2 negative conversion |  |  |
| No. of participants (%) | 12 (100.0%) | 151 (100.0%) |
| Hazard ratio (95% CI) | 0.98 (0.54-1.77) | - |
| log rank P value | 0.946 | - |
| Duration of SARS-CoV-2 positive (day) |  |  |
| Total No. of days | 80 | 917 |
| Mean days of SARS-CoV-2 positive duration | 6.7 | 6.3 |
| P value | 0.724 | - |

There were no significant differences on the percentage of participants with high viral load of these two genes (*p*=1.000 and 0.117, respectively) between the two groups. That is, despite participants received the A8G6 treatment, when they became infected with SARS-CoV-2, they had a comparable level of viral load compared to infected participants in the blank control group (Figure 3). The same analysis conducted in the per protocol set obtained the consistent results.

**Figure 3.**
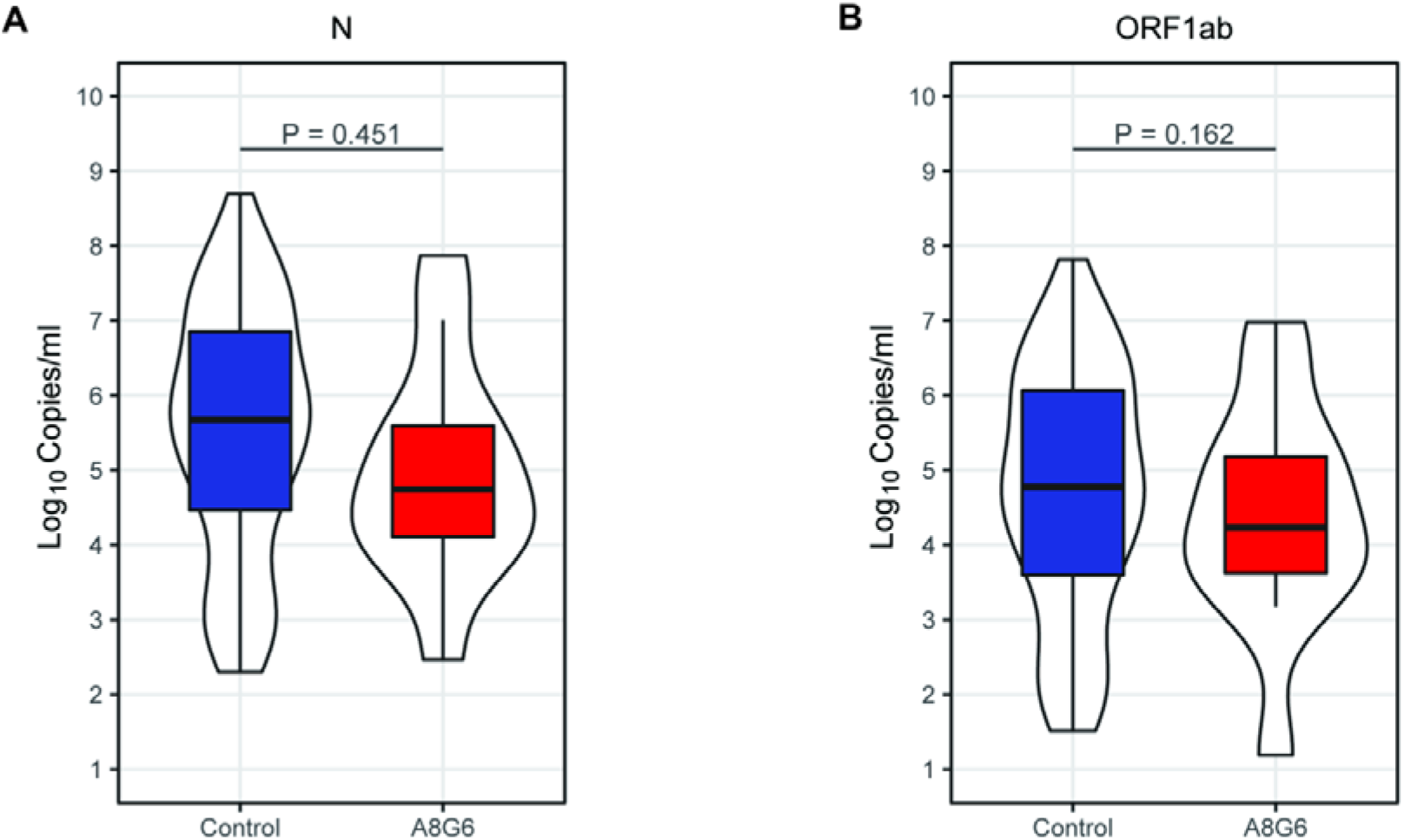
SARS-CoV-2 viral load (log10 copies per ml) at baseline when diagnosed with COVID-19.

### The effect of A8G6 on the time to the COVID-19 recovery

When participants became infected with SARS-CoV-2 in both groups, RT-PCR tests or rapid antigen tests of their oropharyngeal swabs for COVID-19 and the COVID-19 related symptoms were continuously monitored and recorded.

Subjects in both groups who were infected with SARS-CoV-2 during the trial period reported the conversion to COVID-19 negative by the end of the trial. The time of SARS-CoV-2 negativity between groups showed no statistical differences (*p*=0.946) (Figure 4). There is a similar result in the per protocol set.

**Figure 4.**
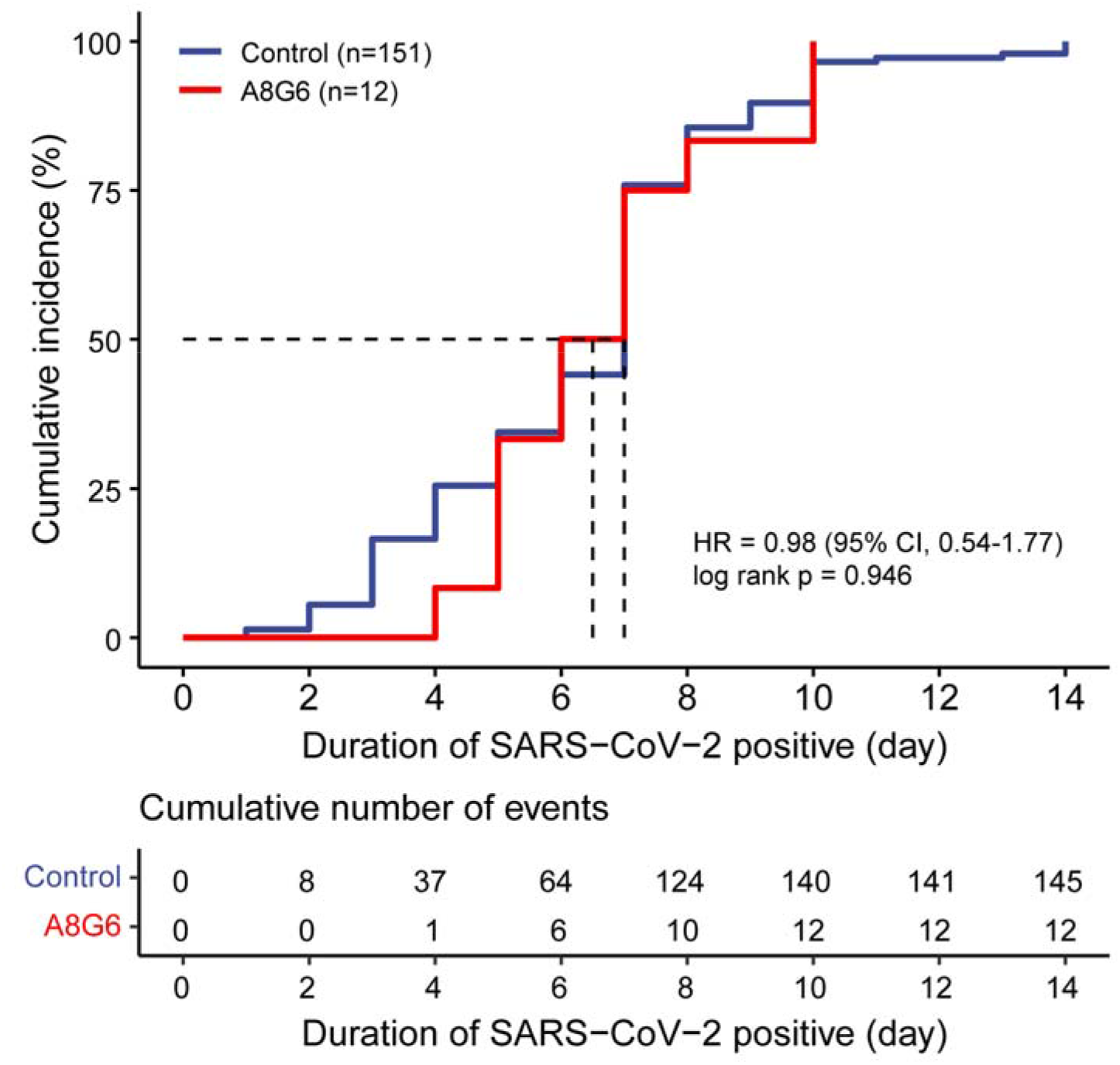
Time-to-event curve for time to viral clearance of SARS-CoV-2.

### Safety

Participants receiving A8G6 treatment (n=173) were required to recorded adverse events (AEs). Thereinto, AEs reported by COVID-19 negative participants (n=161) were not correlated with COVID-19, but might be correlated with the A8G6 treatment. AEs reported by COVID-19 positive participants (n=12) might be correlated with COVID-19 or A8G6. Therefore, after the exclusion of AEs related to COVID-19, the presumptive AEs related to A8G6 treatment were analyzed. Total of 96.9% of the participants in the A8G6 treatment group had no treatment-related adverse effects. Only 3.1% subjects reported one adverse event. The special performance included nasal swelling (N=2, 1.24%), dry throat (N=2, 1.24%) and ageusia (N=1, 0.62%) (Table 4). No adverse events of special interest were reported during the trial period, and no participants withdrew from the trial because of an adverse event. There is a similar result in the per protocol set.

**Table 4.** Individuals (n) having an adverse event (n=161).

| Adverse events | n (%) |
| --- | --- |
| Nasal swelling | 2 (1.24%) |
| Dry throat | 2 (1.24%) |
| Ageusia | 1 (0.62%) |

## 4. Discussion

The nasal spray antibody cocktail A8G6 had demonstrated broad spectrum potency blocking the SARS-CoV-2 infection in our previous preclinical data and also demonstrated favorable safety profile in a first-in-human trial (unpublished data, manuscript in preparation). In this study, we conducted an open-label, non-randomized, two-arm, blank-controlled trial among close contacts of COVID-19 patients in several designated quarantine hotels, during the COVID-19 outbreak occurred in November, 2022 in Chongqing, China.

The intranasal spray antibody cocktail A8G6 was assessed to the effectiveness and safety for the post-exposure prophylaxis of COVID-19 in the real-world. Our data suggest that the application of A8G6 in the close contacts within the 72-hour exposure decreased COVID-19 incidence rate by more than 30%. Moreover, the A8G6 treatment prolonged the occurrence of SARS-CoV-2 infection by at least one day.

At present, most previously authorized COVID-19 antibody treatments under EUA were administrated via vein or intramuscular injection with a high dosage. Those treatment also had several adverse effected that affect quality of life, including pain at the site of injection, allergic reaction, nausea and so on.^15^ As a respiratory pathogen, SARS-CoV-2 infection is primarily caused by breathing in infectious viral particles through nasal airway. An intranasal spray of neutralizing antibodies may provide a more direct protection against viral entry.

Moreover, this non-invasive drug delivery is easier to use and may result in better medication compliance. In our study, the favorable safety profile of A8G6 with the few adverse effects was consistent with other nasal spray drugs.^16^ Thus, A8G6 can be used in a wide range of population, especially in some special population with comorbidities and immunocompromised population. The effective treatment of A8G6 among high-risk patients could reduce medical cost, usage of medical resources and COVID-19 transmission risk. Furthermore, participants who experienced SARS-CoV-2 infection under the A8G6 treatment, showed delayed COVID-19 infection by ∼ 1 day, which could provide important relieve on medical resources at the epidemic peak.

Currently, there were a few other antibody nasal sprays in clinical development. The neutralization efficacy of nasal mucosal samples against SARS-CoV-2 variants after the nasal spray treatment of a monoclonal antibody 35B5 was calculated as 60%.^17^ The effectiveness of the SA58 nasal spray was evaluated as 77.7% (95% CI: 52.2% - 89.6%) and 61.83% (95% CI: 37.5% - 76.69%) in medical personnel and healthy workers, respectively.^18, 19^ In our primary endpoint analysis, the nasal spray A8G6 antibody cocktail showed decreased risk of infection of close contacts with COVID-19 patients. The prevention efficacy of the A8G6 treatment within 72-hour exposure was calculated to be 84.4% (95% CI: 74.4-90.4). A8G6 showed comparable or better COVID-19 prevention in the real world than other similar antibody nasal spray.

Current data in this study showed that 6.9% of A8G6 treated participant became SARS-CoV-2 positive (vs 44.4% in the blank control group) during the study period. Our results suggested that post-infection A8G6 treatment provided limited benefits on viral load reduction and time to viral clearance. This is consistent with the potential mechanism of action of A8G6 nasal spray. Once SARS-CoV-2 virus enters into the cells and starts viral replication, A8G6 neutralizing antibody has limited efficacy to stop the viral replication. Our data also indicated that baseline characteristics of these two groups in the efficiency of viral replication were similar.^20^ In another study, the similar viral load were also reported between the vaccinated individuals with breakthrough infections and unvaccinated individuals with SARS-CoV-2 infection.^21^

There were several limitations for this study. First limitation is the lack of a placebo arm. We did not conduct this study with the double-blind procedure because there was a small window of time to initiate and complete the study so not allowing enough time for the placebo to be produced before the trial.

Second limitation is the lack of participant randomization in the study design. This was primarily due to a large percentage of eligible participants not feel comfortable to take the A8G6 treatment at time of enrollment. Therefore, we have to assign those participants to blank-controlled group. Under this situation, complete randomization was not possible. Third limitation is the lack of participants developing severe COVID-19 that need hospitalization due to small sample size. Therefore, this study did not assess the efficacy of A8G6 in preventing severe COVID-19. During the study period, there was an adjustment of the public health policy of the COVID-19 pandemic in China, that the SARS-CoV-2 infected persons no longer were reported in the future. As a result, the definition of close contacts became difficult and it became difficult to enroll more participants to increase the sample size. Fourth limitation is that the study was conducted in the designated quarantine hotels. Study participants were assumed to be single-exposure to positive COVID-19 individuals. The effects of increased infection risks of multiple exposures in the real world on the A8G6 efficacy should be considered in the further study.

In conclusion, we observed potent post-exposure prevention efficacy of intranasal spray AG86 antibody combination in close contacts of COVID-19 patients. This proof-of-concept study result suggested the potential beneficial effect of neutralizing antibody administrated as nasal spray in COVID-19 prevention. Currently A8G6 nasal spray is under clinical development to further assess its efficacy and safety.

## Contributors

AH, NT and XL designed the trial and study protocol. PP contributed to the literature search. XL and HD verified the data. PP and HD wrote the first draft manuscript. AH, NT, XL, PP, HD and ZY contributed to the data interpretation and revision of the manuscript. HD, XL, NT and PP contributed to data analysis. XL monitored the trial. QY, SC, BL, MH and XL were responsible for the site work including the recruitment, follow-up, and data collection, and XL was the site coordinator. All the authors had full access to all the data in the study and had final responsibility for the decision to submit for publication.

## Data Availability

All data produced in the present study are available upon reasonable request to the authors.

## Declaration of interests

All authors declare no competing interests.

## Data sharing

De-identified individual participant-level data will be available upon written request to the corresponding author following publication.

## Acknowledgements

We thank Dr. Yang Tian and Chengyong Yang (Mindao Haoyue Co., Ltd., Chongqing, China) for the constructive suggestion about the trial design and manuscript. We thank all the participants who took part and contribute specimens in our study. We also thank the support from Yuzhong District Center for Disease Control and Prevention (Chongqing) and all medical personnels who worked hard in this trial.

## Notes

### Competing Interest Statement

Ailong Huang declares the following competing interests: Patent has been filed for 
some of the antibodies presented here (patent application number: PCT/CN2020/115480, 
PCT/CN2021/078150, PCT/CN2021/113261; patent applicants: Chongqing Medical University). 
All other authors declare no competing interests.

### Clinical Trial

ChiCTR2200066416

### Funding Statement

This study was funded by Chongqing Biomedical R&D Major Special Project, Project (No. CSTB2022TIAD-STX0013), Chongqing Science and Health Joint Medical High-end Talent Project (No. 2022GDRC012), Science and Technology Research Program of Chongqing Municipal Education Commission (No. KJZD-K202100402), CQMU Program for Youth Innovation in Future Medicine (No. W0073).

### Author Declarations

The protocol has been approved by the Ethics Committees of The Second Affiliated Hospital of Chongqing Medical University (the approval number: 2022127-1).

